# Predicting Dementia in People with Parkinson’s Disease

**DOI:** 10.1101/2025.01.27.25321134

**Authors:** Mohamed Aborageh, Tom Hähnel, Patricia Martins Conde, Jochen Klucken, Holger Fröhlich

## Abstract

Parkinson’s disease (PD) exhibits a variety of symptoms, with approximately 25% of patients experiencing mild cognitive impairment and 45% developing dementia within ten years of diagnosis. Predicting this progression and identifying its causes remains challenging. Our study utilizes machine learning and multimodal data from the UK Biobank to explore the predictability of Parkinson’s dementia (PDD) post-diagnosis, further validated by data from the Parkinson’s Progression Markers Initiative (PPMI) cohort. Using Shapley Additive Explanation (SHAP) and Bayesian Network structure learning, we analyzed interactions among genetic predisposition, comorbidities, lifestyle, and environmental factors. We concluded that genetic predisposition is the dominant factor, with significant influence from comorbidities. Additionally, we employed Mendelian randomization (MR) to establish potential causal links between hypertension, type 2 diabetes, and PDD, suggesting that managing blood pressure and glucose levels in Parkinson’s patients may serve as a preventive strategy. This study identifies risk factors for PDD and proposes avenues for prevention.

## 2 Introduction

Parkinson’s disease (PD) is the fastest growing neurological disease and the second most common neurodegenerative disease [1]. Although the cause of PD remains unclear in most sporadic cases, mutations in more than 20 genes have been associated with the disease, including genes such as LRRK2 and SNCA [2]. In addition, environmental risk factors have been linked to the development of PD, which can be triggered by an underlying genetic predisposition [3]. For example, occupational exposures such as pesticides are associated with an increased risk of PD [4][5][6]. Furthermore, lifestyle factors such as alcohol consumption, physical inactivity and diet [7], as well as beta-blockers [8] and comorbidities such as diabetes mellitus and arterial hypertension, have been identified as potential risk factors.

Although PD is characterized primarily by three main motor symptoms (bradykinesia, tremor at rest, and rigidity); cognitive impairment is also a frequent problem in patients with PD (PwPD). Approximately 25% of PwPD exhibit mild cognitive impairment and 45% develop dementia within 10 years after diagnosis [9][10][11][12][13][14]. PD dementia (PDD) contributes to an increase in health-related expenditures and a significant decrease in quality of life, underscoring its importance to affected individuals, their caregivers and healthcare systems [15]. Furthermore, PDD constitutes one of the four milestones that occur on average four years before death, which indicates the onset of the terminal phase of the disease [16]. The underlying reason why certain PwPD develop cognitive impairment earlier in their disease course is still unknown. Previous genome-wide association studies (GWAS) report possible associations of different genetic risk variants with cognitive impairment in PD, including variants in SNCA [17][18], MAPT [19][20], TMEM175 [21] and GBA [22][23]. Statistical analysis of a cohort of 827 individuals identified age, duration of the disease, sex, and GBA status as the primary factors associated with cognitive performance and progression to dementia [24].

[25] developed machine learning models to predict cognitive outcomes in de novo diagnosed PwPD using a broad spectrum of baseline data (including cognitive test results) from the Parkinson’s Progression Markers Initiative (PPMI) cohort study. However, data from specific cohorts are not necessarily representative of the overall disease population and could therefore differ from real-world data collected in population studies or clinical routine. Moreover, models trained on data collected solely for research, such as DNA methylation, cerebral spinal fluid biomarkers and neuropsychiatric tests, are not easily applicable in clinical practice. Thus, there is a need to investigate models using more routinely collected health data, including comorbidities, lifestyle and environmental factors. The potential impact of such factors on PDD is important for shaping future prevention strategies.

Our study used the UK Biobank (UKB) to a) predict dementia in people with Parkinson’s disease (PwPD) using machine learning and b) investigate modifiable risk factors that can causally influence the development of dementia in PwPD, thus informing possible prevention strategies. To our knowledge, no prior research has explored these areas.

To achieve our objectives, we developed and evaluated machine learning models to predict PDD based on comorbidities, genetic, environmental, and lifestyle factors. Using Explainable AI (XAI) techniques, we identified the features that significantly affected predictions and used probabilistic graphical models (Bayesian networks) to analyze interactions among the most predictive comorbidities, environmental, lifestyle, and genetic risk factors. Finally, we performed a Mendelian Randomization (MR) analysis to confirm the probable causal relationship between hypertension, type 2 diabetes, and PDD.

## 3 Methods

### 3.1 Data Sources

The UKB is a large-scale database and research resource, playing an important role in studying a wide spectrum of complex diseases and advancing their diagnosis, prevention, and treatment [26]. The cohort was released for research in April 2012, and currently includes cross-sectional data for more than 500,000 individuals from the UK. The initial assessments involved the collection of data on demographics, clinical history, and lifestyle. The participating individuals were also genotyped, providing valuable genetic data that can facilitate the study of the genetic architecture of complex diseases.

To further validate our findings from UKB, we used data from the PPMI study, which provides multiple data modalities, including demographic and genetic components [27]. The data set was initially released in 2010, and currently includes data for more than 1500 patients.

### 3.2 Patients Selection and Variables

We selected all UKB patients diagnosed with PD in the hospital according to the ICD-10 code G20 and those with Parkinson’s Dementia (F02.3). This resulted in 3,541 PwPD and 487 patients with PDD. Data were cross-sectional and included past diagnoses, comorbidities, as well as genetic, environmental, and lifestyle factors associated with the risk of PD, as identified in a large meta-analysis [28]. Following this publication, we constructed variables that reflect known risk factors associated with dementia [29], including stroke, type 2 diabetes, hypertension, depression, anxiety, hearing loss, visual loss, hypercholesterolemia and obesity. For daytime sleeping/dozing, we used measurements from the UKB field 1220 (daytime dozing / sleeping). All variables are detailed in Supplementary Table 1.

For PPMI, we included PwPD and healthy controls, accounting for their cognitive status to include individuals with mild cognitive impairment (MCI) and dementia as recorded in the PPMI cognitive state table (code 2 indicating MCI and code 3 indicating dementia). Together, the data set included a total of 637 PwPD, among which 122 had PDD.

### 3.3 Use of Genotype Data

Genotyping was performed on all UKB participants using UKBiLEVE and UK Biobank Axiom arrays. The UKBiLEVE array was initially designed and ran on approximately 50,000 participants, which was followed by the UK Biobank Axiom array designed to run on the remaining 450,000 participants. Both arrays share over 95% common content. For PPMI, more than 1500 participants were genotyped using the Illumina NeuroX array, which combines approximately 240,000 exome variants and approximately 24,000 custom variants with a focus on neurological diseases.

Since PDD is defined as dementia due to PD, we hypothesized that certain genetic variants associated with PD could also contribute to the risk of PDD. To construct variables for our machine learning models, we thus obtained 3189 single nucleotide polymorphisms (SNPs) associated with PD from the ieu-b-7 dataset. In addition, variants associated with all-cause dementia might play a role. Hence, we obtained 924 dementia-associated SNPs from the finn-b-F5 Dementia dataset available on the ieugwas database (8 April 2024) [30]. We performed SNP clumping to include only highly disease-associated, rather independent SNPs, using thresholds of *p* < 5*E* − 8, *r*^2^ < 0.1 and a physical distance of 250 kb. This process resulted in 10 highly associated SNPs for PD and 13 for dementia. In addition, we calculated two polygenic risk scores that are published in the Polygenic Risk Score Catalog (PGS) [31], namely PGS4281 for dementia [32] and PGS777 for PDD [33]. Finally, we included 3 SNPs that have been associated with PDD from the DisGeNet gene-disease association network [34].

### 3.4 Machine Learning Models Predicting PD Dementia

We built three classification models to predict PDD within PwPD. This included gradient boosting [35], random forests [36] and elastic net penalized logistic regression [37]. The aim was to predict PDD using multimodal data, including SNP, polygenic risk scores, demographic characteristics, comorbidities, family history, and environmental and lifestyle-associated factors. The models were trained within a 5-fold nested cross-validation approach which splits data into a training set (80%) and a testing set (20%) while optimizing the model’s hyperparameters within an inner cross-validation on the training set. Details on hyper-parameter optimization are presented in Supplementary Table 2. We evaluated the models’ predictive performance by calculating the area under the receiver operating characteristic curve (AUC) across 20 repetitions of 5-fold nested cross-validation. To address label imbalance, class weights were applied during model training.

### 3.5 Making Model Predictions Explainable

We calculated Shapley Additive Explanations (SHAP values) from the best performing machine learning model to understand the importance of features and the cumulative influence of different data modalities on predictions [38].

To better understand the interactions between all variables, we subsequently learned a Bayesian network (BN) from the data [39]. BNs are probabilistic graphical models that represent variables as nodes and conditional probabilistic dependencies between them as edges. We trained a BN on data from all subjects using non-parametric bootstrapping, which randomly selects samples 1000 times and learns a complete BN graph structure within each bootstrap using the tabu metaheuristic search method, which we have previously found to be the most accurate compared to multiple other BN structure learning methods for multimodal clinical data [40]. Continuous variables were discretized through clustering of k-means before learning the BN structure to account for the non-Gaussian nature of multiple features. BN structure learning was implemented using R-package bnlearn [41]. As each bootstrap sample resulted in a slightly different BN structure, a consensus structure was subsequently created by applying a conservative bootstrap probability threshold of 0.5, meaning that the edges of the averaged graph were found in at least 50% of all BN. This aims to improve the robustness and reliability of the estimates. Further details of BN learning can be found in Supplementary Table 3.

### 3.6 Mendelian Randomization to Estimate Causal Effects

MR assesses the causal relationship between an exposure (here: a comorbidity) and an outcome (here: PDD) using genetic variants as instrumental variables (IV). The variants used should satisfy three assumptions to be considered as an IV:

- The variant is associated with the exposure
- The variant is independent of confounding factors that confound the association of the exposure to the outcome
- The variant is independent of the outcome given the exposure and the confounding factors

In this regard, the genetic variants associated with exposure can be used as proxies to explain how exposure can influence the outcome of interest. For our study, we used MR to assess the putative causal effect of comorbidities on the risk of PDD.

We obtained genetic variants for our exposures using the IEU-OpenGWAS API (ieugwasr package [42]) from summary datasets for stroke (ebi-a-GCST005838), type-2 diabetes (ebi-a-GCST90018926), hypertension (ebi-a-GCST90038604), depression (ukb-b-12064), anxiety (ukb-a-82), hypercholesterolemia (finn-b-E4 HYPERCHOL), hearing loss (finn-b-H8 CONSENHEARINGLOSS), visual loss (finn-b-H7 VISUALDISTBLIND), obesity (finn-b-E4 OBESITY), smoking status (ebi-a-GCST90029014), and daytime sleeping/dozing (ukb-b-5776). The selection of summary datasets was based on the availability of top hits. We conducted one-sample MR by assessing the associations between genetic variants and both the exposures and the outcome at the individual level. We calibrated SNP exposure estimates to our phenotypes and generated SNP outcome estimates through association testing, adjusting for age and sex. The instruments were selected by clumping to ensure that the SNPs were strongly associated with exposures and independent of each other, applying thresholds of *p* < 5*E* − 8, *r*^2^ < 0.1 for linkage disequilibrium and a genomic window of 250 kb. We estimated causal effects using the inverse variance weighted method (IVW) with fixed effects, which calculates the causal effect of exposure *X* on outcome *Y*. The causal effect ratio using a genetic variant *i* is *X*_*i*_*/Y*_*i*_, with the standard error estimated by the delta method as 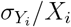.The IVW estimate combines ratio estimates of the variants in a fixed-effect meta-analysis model.

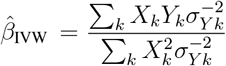

The success of MR depends on the three previously mentioned assumptions, which require a sensitivity analysis for the evaluation of the results [43]. We utilized MR-Egger and MR-PRESSO for this analysis. The MR-Egger method evaluates directional pleiotropy, assessing whether genetic factors have average pleiotropic effects on the outcome that differ from zero. Pleiotropy in human genetics can manifest in various forms, such as a single variant influencing multiple traits or a causal locus affecting a trait through another one. Horizontal pleiotropy occurs when a variant directly or indirectly impacts the target outcome, often by influencing other traits that causally affect it. In this context, MR-Egger provides a reliable estimate of the causal effect under the assumption of InSIDE (Instrument Strength Independent of Direct Effect) [44]. In this work, MR-PRESSO was used to investigate horizontal pleiotropy in multi-instrument summary level MR [45], which involves three steps: a global test for horizontal pleiotropy detection, removal of outliers for correction, and a significance test before and after removal of outliers.

## 4 Results

### 4.1 Demographics of Patients and Controls

In UKB, PwPD were more likely to be male (61.3%) with a mean age of 62.7 years, consistent with previously reported incidences [46][47]. The PDD group showed an even higher proportion of males (69.8%) and a slightly higher mean age of 64.1 years.

In PPMI, PwPD showed a proportion of male individuals similar to UKB (60.4%) with a mean age of 61.8 years. The PDD group showed a slightly lower proportion of male individuals than in UKB (61.5%), and the mean age was 67.1 years. Detailed demographic characteristics of the groups can be found in Table 1.

**Table 1:**
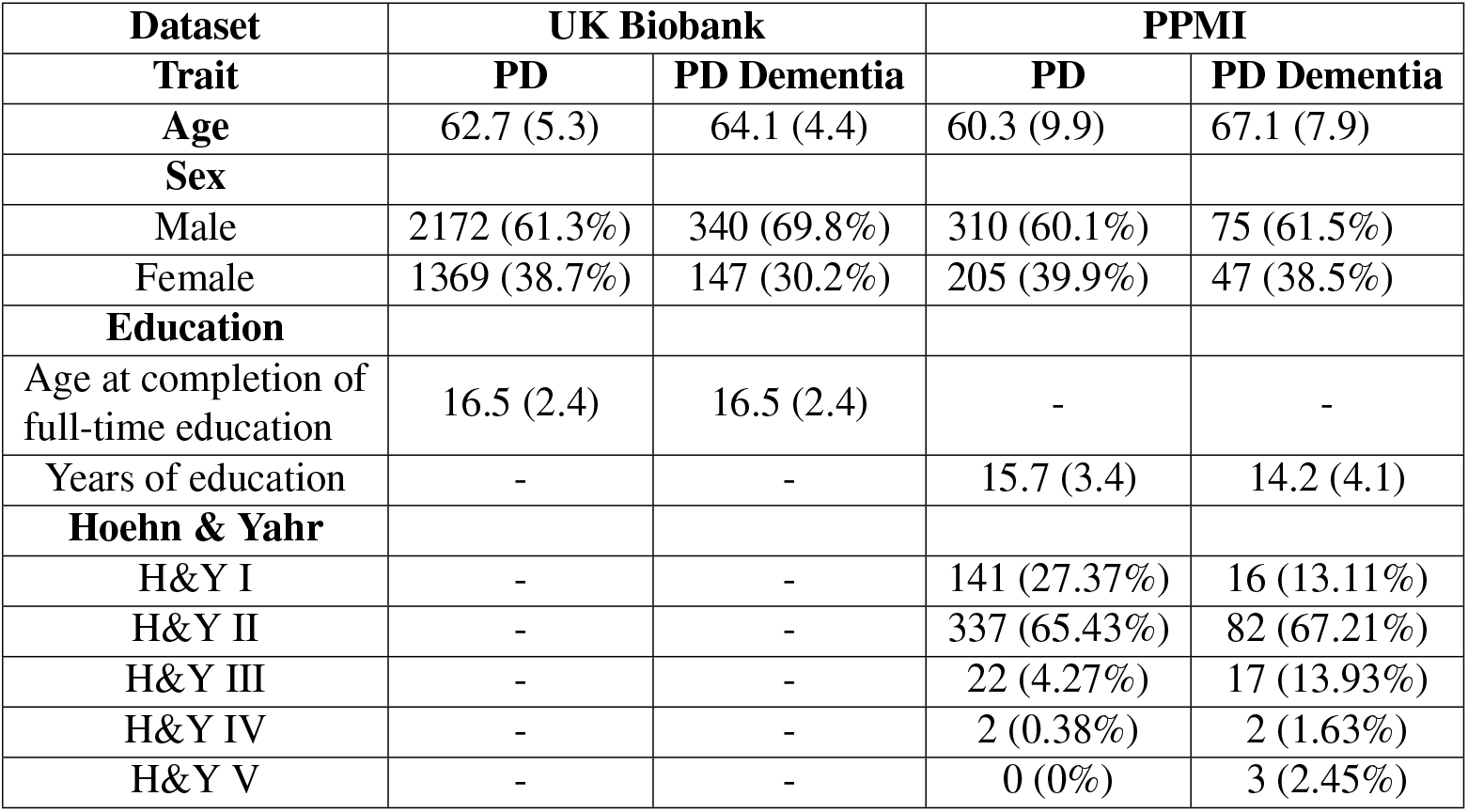
Demographic characteristics of Parkinson’s disease (PD) and Parkinson’s disease dementia (PDD) groups in the UK Biobank and PPMI datasets. Continuous variables are reported as mean (standard deviation), while categorical variables are presented as count (percentage).

| Dataset | UK Biobank |  | PPMI |  |
| --- | --- | --- | --- | --- |
| Trait | PD | PD Dementia | PD | PD Dementia |
| Age | 62.7 (5.3) | 64.1 (4.4) | 60.3 (9.9) | 67.1 (7.9) |
| Sex |  |  |  |  |
| Male | 2172 (61.3%) | 340 (69.8%) | 310 (60.1%) | 75 (61.5%) |
| Female | 1369 (38.7%) | 147 (30.2%) | 205 (39.9%) | 47 (38.5%) |
| Education |  |  |  |  |
| Age at completion of full-time education | 16.5 (2.4) | 16.5 (2.4) | - | - |
| Years of education | - | - | 15.7 (3.4) | 14.2 (4.1) |
| Hoehn & Yahr |  |  |  |  |
| H&Y I | - | - | 141 (27.37%) | 16 (13.11%) |
| H&Y II | - | - | 337 (65.43%) | 82 (67.21%) |
| H&Y III | - | - | 22 (4.27%) | 17 (13.93%) |
| H&Y IV | - | - | 2 (0.38%) | 2 (1.63%) |
| H&Y V | - | - | 0 (0%) | 3 (2.45%) |

### 4.2 Predictability of PD Dementia

We evaluated penalized logistic regression, Random Forests, and XGBoost to predict PDD. Repetitive nested cross-validation showed an average AUC of 0.62 for Random Forest and logistic regression and 0.61 for XGBoost over 20 repetitions (Figures 1 and 2). We carried out an ablation study to understand the contribution of different data modalities. We found that demographics, comorbidities, and genetics contributed the most, with a statistically significant average of 2% drop in AUC after removing any of them; see Tables 2, 3.

**Figure 1:**
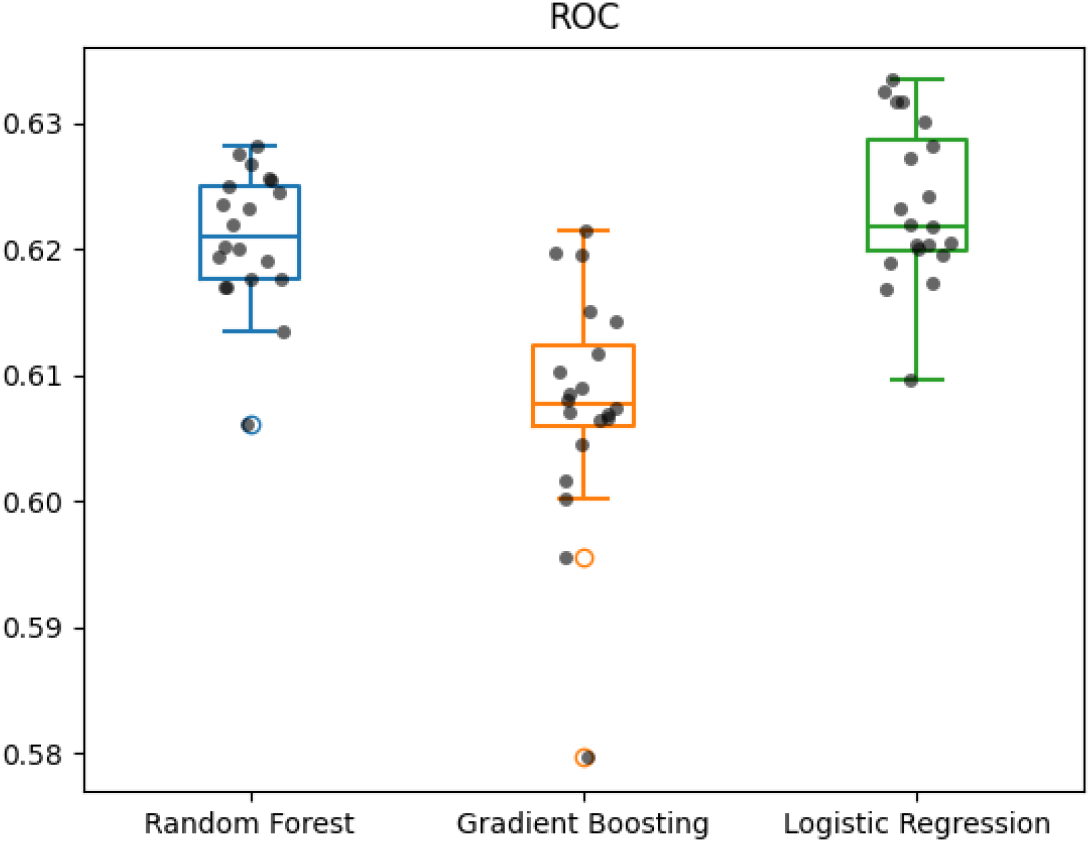
Boxplot showing the models’ AUC via repeated nested cross-validation

**Figure 2:**
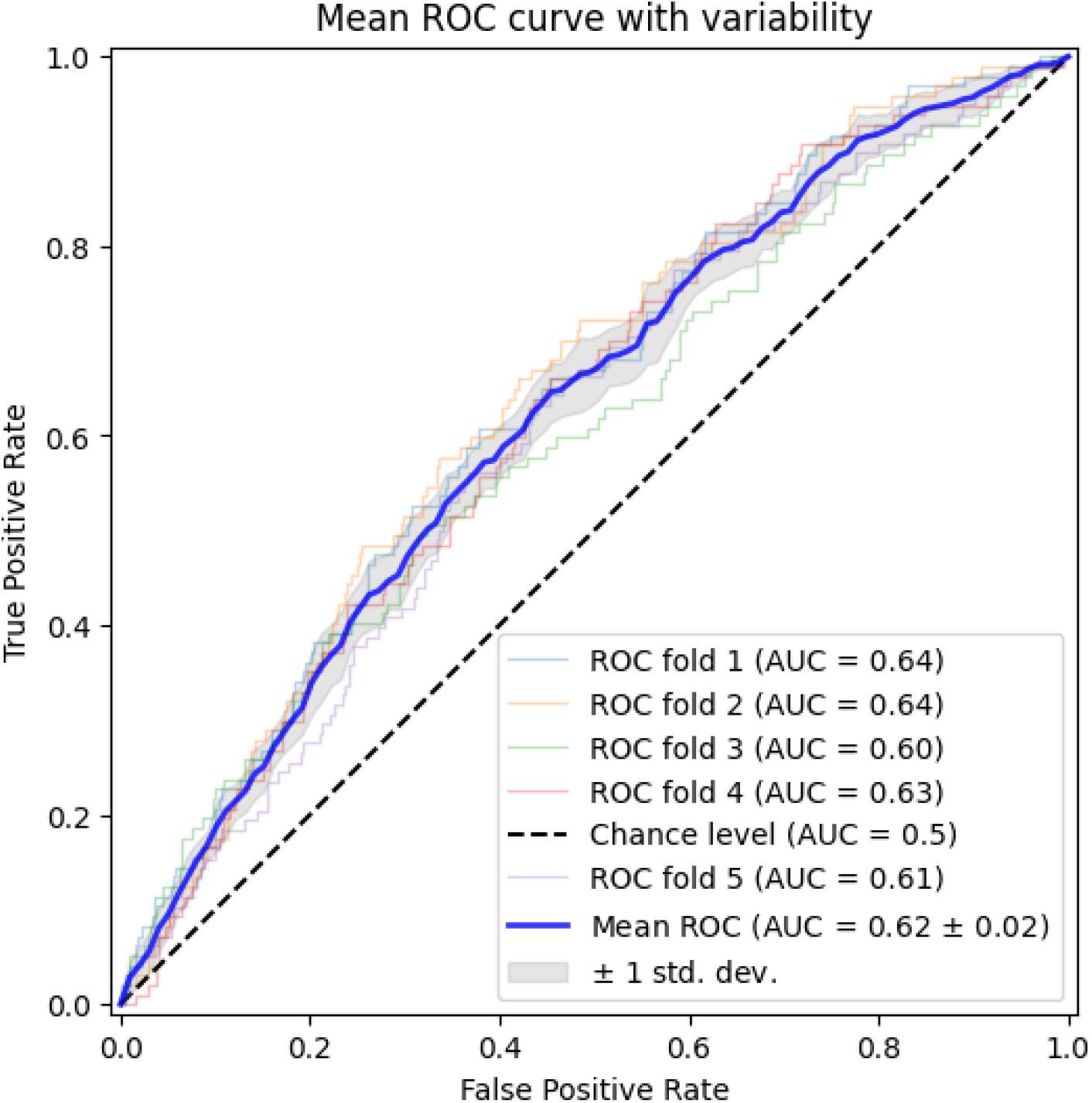
Variability of mean ROC curve of the Random Forest model for each fold via cross validation

**Table 2:** Comparison of the model’s performance showing the median AUC over 20 repeats of nested cross-validation, using all data modalities and each modality separately. A Kruskal-Wallis test was used to determine if there were statistically significant differences in performance after the removal of each modality compared to the full model.

| Modality | Random Forest |  | XGBoost |  | Logistic Regression |  |
| --- | --- | --- | --- | --- | --- | --- |
|  | Median AUC | P-value | Median AUC | P-value | Median AUC | P-value |
| Multimodal | 0.62 (0.005) | - | 0.60 (0.009) | - | 0.62 (0.006) | - |
| Demographics + Genetics | 0.61 (0.004) | <0.05 | 0.59 (0.007) | <0.05 | 0.61 (0.006) | <0.05 |
| Demographics | 0.56 (0.005) | <0.05 | 0.54 (0.008) | <0.05 | 0.57 (0.004) | <0.05 |
| Genetics | 0.58 (0.003) | <0.05 | 0.56 (0.008) | <0.05 | 0.58 (0.05) | <0.05 |
| Comorbidities | 0.58 (0.004) | <0.05 | 0.57 (0.004) | <0.05 | 0.57 (0.03) | <0.05 |
| Lifestyle | 0.49 (0.01) | <0.05 | 0.5 (0.009) | <0.05 | 0.49 (0.005) | <0.05 |
| Environmental | 0.51 (0.007) | <0.05 | 0.51 (0.008) | <0.05 | 0.5 (0.006) | <0.05 |
| Family history | 0.49 (0.01) | <0.05 | 0.49 (0.01) | <0.05 | 0.49 (0.006) | <0.05 |

**Table 3:**
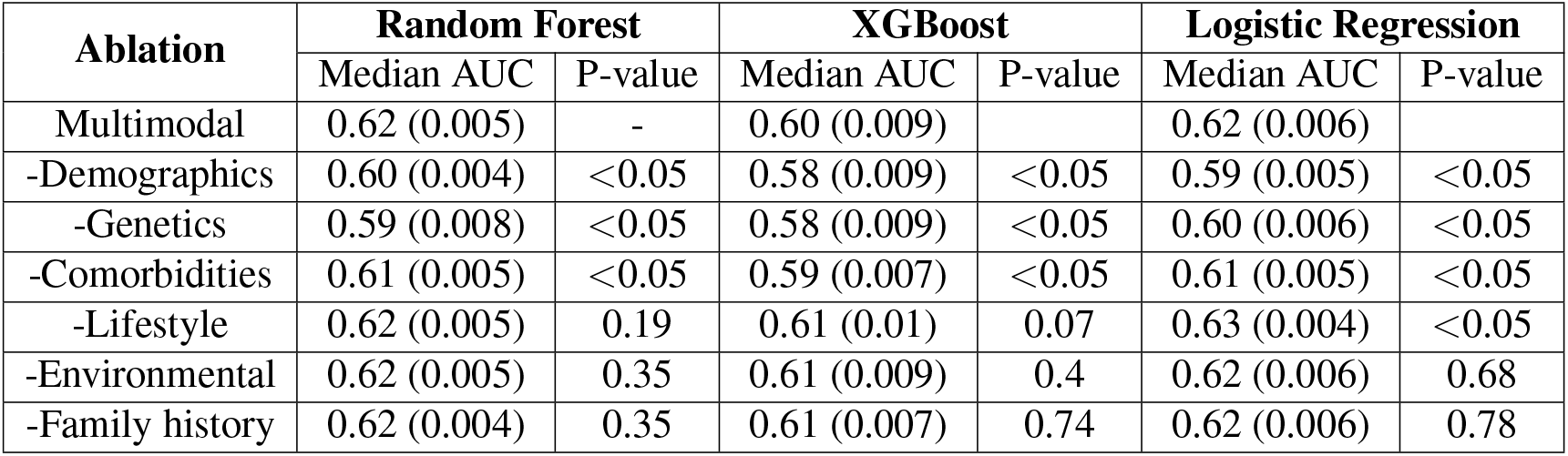
Ablation study results showing the median AUC over 20 repeats of nested crossvalidation after removing each data modality. A Kruskal-Wallis test was used to determine if there were statistically significant differences in performance after the removal of each modality relative to the full model.

We repeated our analysis using the PPMI dataset and compared the results with those obtained from the UKB cohort. Due to the absence of several variables in the PPMI dataset that were only available in UKB, we had to take a reduced subset of variables. This subset included only SNPs, PRS, age, and sex. When training the Random Forest model on UKB with this reduced subset of variables, the cross-validated prediction performance of this reduced model was close to the original with an average AUC 0.61 *±* 0.01, while the cross-validated prediction performance on PPMI was higher with an average AUC 0.65 *±* 0.02 (Figure 3).

**Figure 3:**
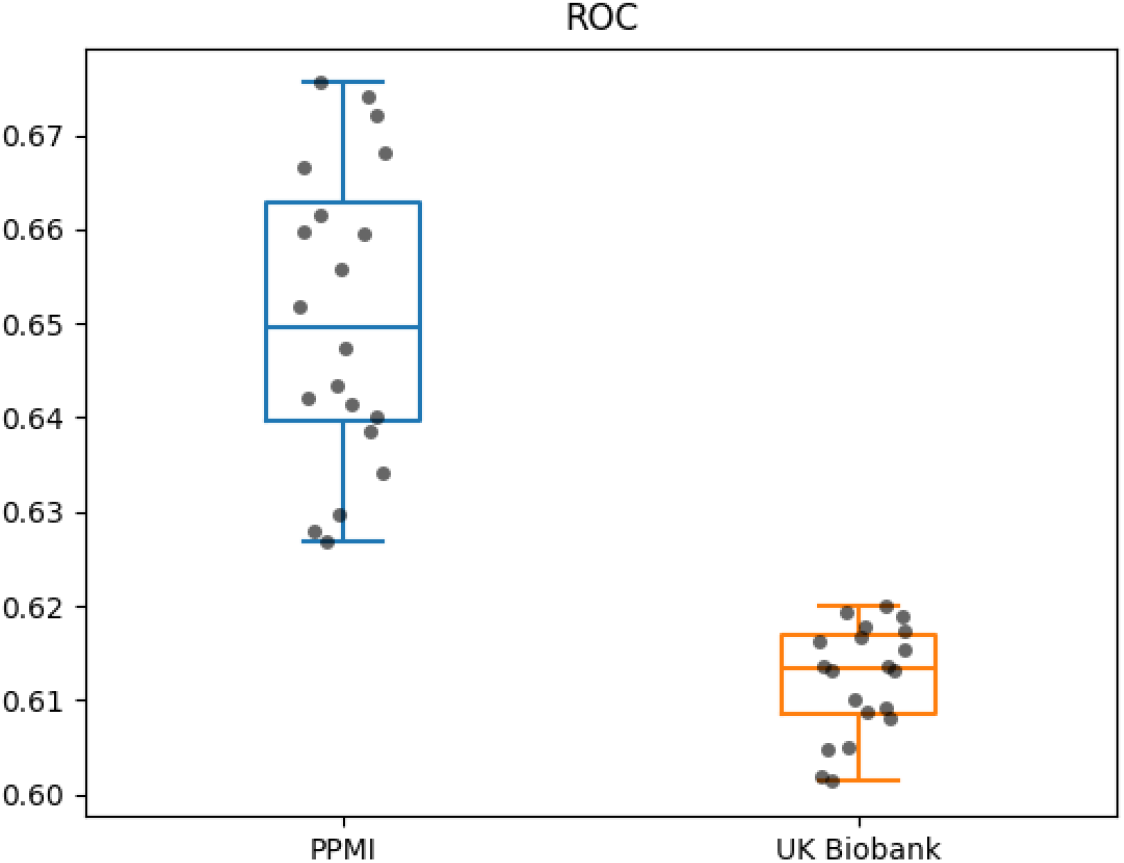
Comparison of the Random Forest model trained on the reduced subset of features in PPMI and UK Biobank datasets

### 4.3 SHAP Analysis

For the following analysis, we focused on the best performing model variant after training and tuning on the entire dataset, i.e. the Random Forest model using all data modalities.

SHAP analysis revealed that variables from different data modalities contribute significantly to the predictions of the model, with the polygenic risk score PGS4281 being the main predictor, followed by SNP *rs769449*, age, SNP *rs6859*, diagnosis of depression, sex, BMI and hypercholesterolemia. We also observed that lifestyle variables, such as tea intake and water intake, appeared among the top predictors. Figure 4 shows the impact of the top 20 variables on the model output and Figure 5 shows the relationship between variable values and model predictions using SHAP dependence. We found that higher PGS4281 scores and older age were associated with higher SHAP values, indicating a stronger influence on the predicted probability of PDD. In contrast, a higher BMI was associated with lower SHAP values, i.e. lower predicted likelihood of PDD. For SNP *rs769449*, having one or two copies of the effect allele resulted in higher SHAP values. In contrast, for SNPs *rs6859* and *rs449647*, having more copies of the effect allele was associated with lower SHAP values. Furthermore, a higher predicted likelihood of PDD was associated with being male and having a diagnosis of depression.

**Figure 4:**
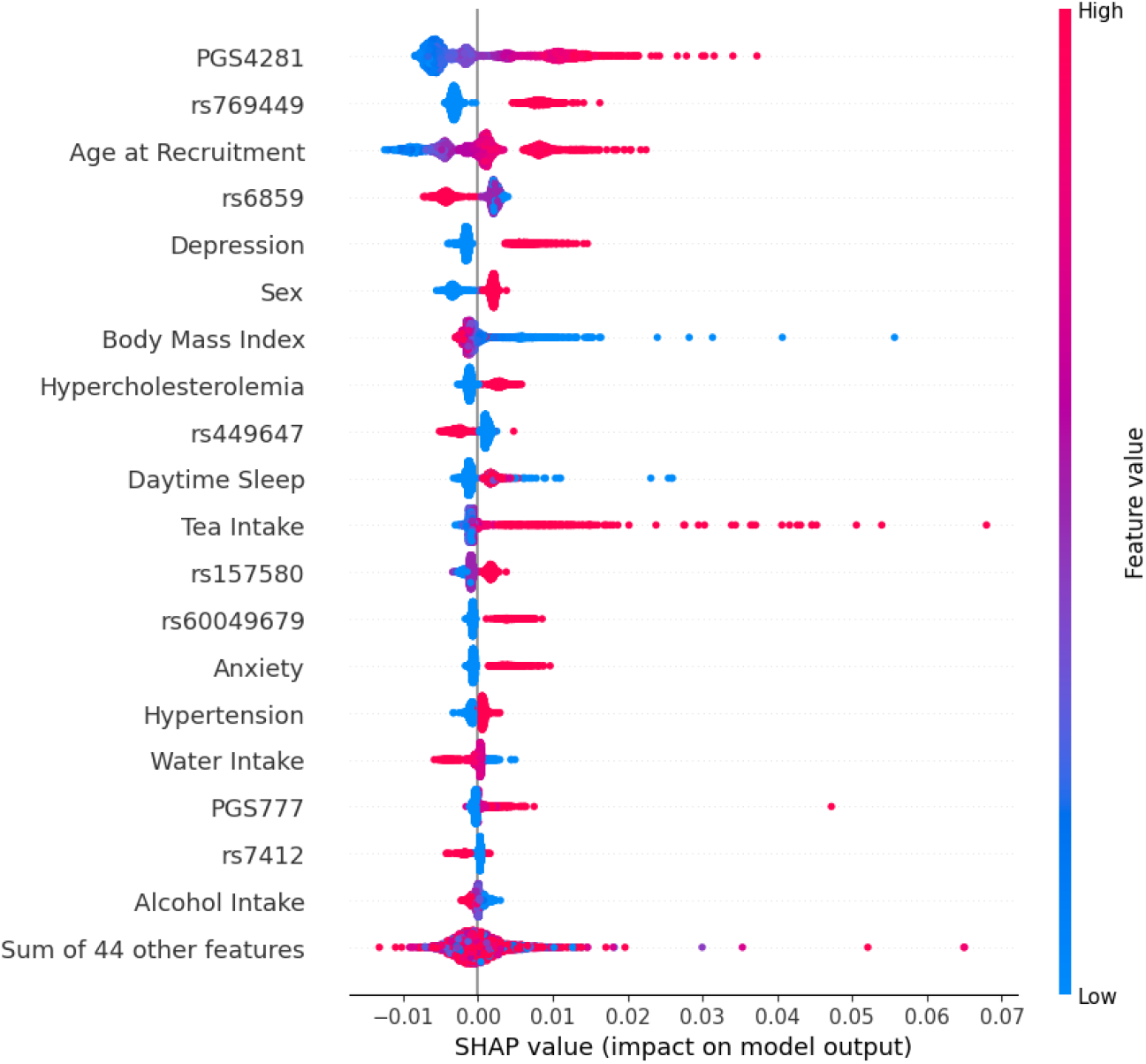
Beeswarm plot showing the SHAP values of the top level predictors for the best performing random forest model

**Figure 5:**
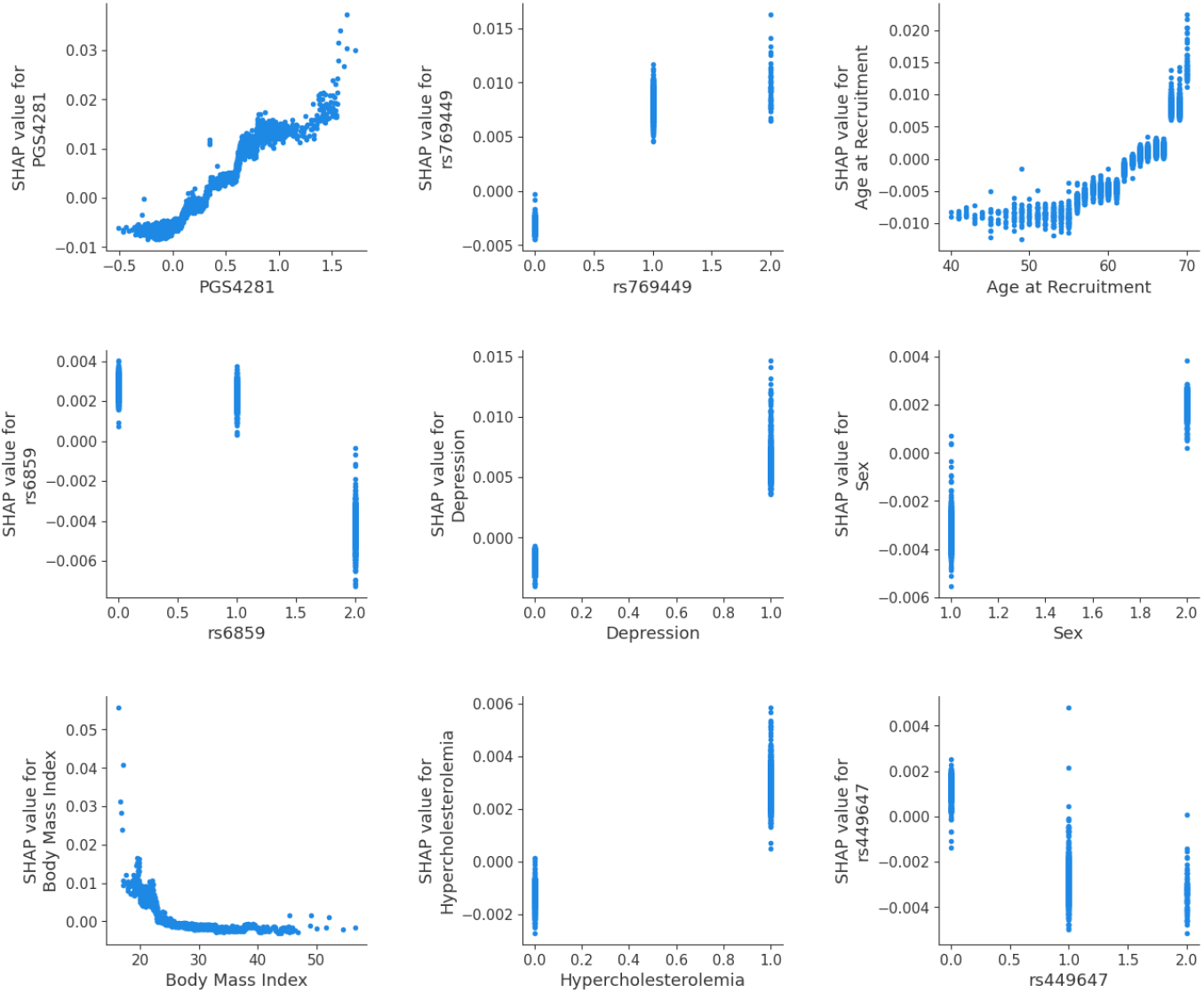
A grid showing the SHAP dependence plot of the top 9 features

Finally, we analyzed the cumulative influence of different data modalities by summing the SHAP values of their respective variables to understand their overall contribution to model predictions. In agreement with the previously presented ablation study, we found that genetics had the highest influence (49.31%), followed by demographics (24.32%) and comorbidities (15.74%). Figure 6 presents the cumulative influence of all data modalities.

**Figure 6:**
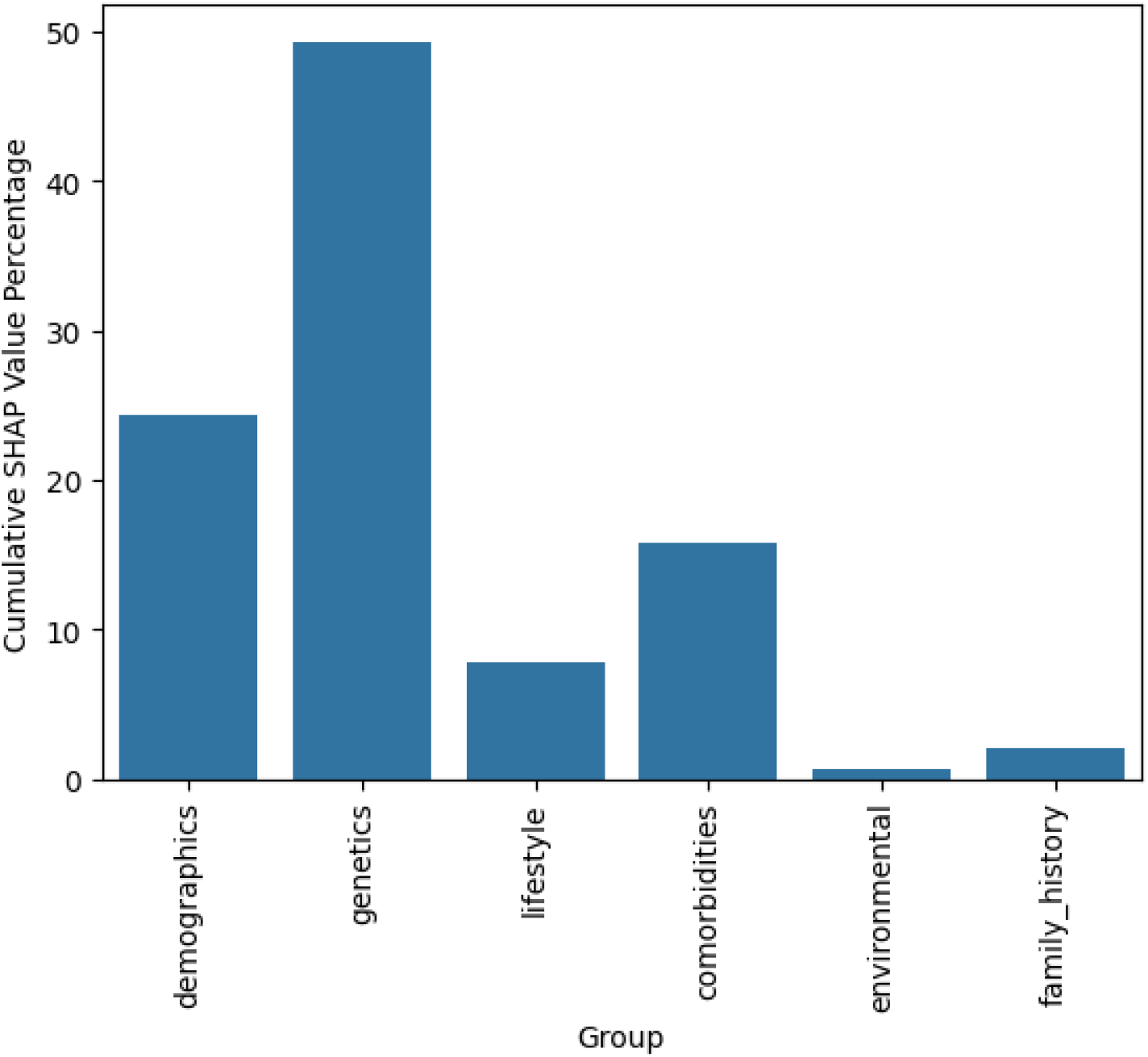
A bar plot illustrating the cumulative influence of different data modalities, with values normalized to a range between 0 and 1 to facilitate comparison.

The SHAP analysis of the Random Forest trained on the entire PPMI data set showed similar results with age as the main predictor, followed by PGS4281, SNP *rs2927468, rs449647* and *rs356219*, while sex and *rs769449* had lower SHAP values. A comparison of the beeswarm plots for both datasets, highlighting the SHAP values of the top predictors, is provided in Supplementary Figure 1.

### 4.4 Understanding the Interactions of Predictors

Following our SHAP analysis, we fitted 1000 BNs within a bootstrap procedure to better understand conditional statistical dependencies between all variables in the best performing Random Forest model trained on UKB. All edges discussed below had a bootstrap frequency of (≥ 50%), which means that they were observed in at least 50% of the BNs across all bootstrap samples; thus, these edges had high statistical confidence. A visualization of these high confidence edges as a graph is provided in Supplementary Figure 2.

Overall, our analysis revealed expected connections within each data modality, including edges that point from diabetes and obesity to hypertension and hypercholesterolemia, from age to anxiety (which also had an edge toward depression), and from sex to diabetes, smoking, anxiety, hypercholesterolemia, and breastfed. In addition, we observed connections among different SNP groups, including SNPs that map to the genes *TMEM175, NSF, LRRC37A2, SNCA* and *KANSL1*. We also observed connections between genetic and non-genetic variables, namely an edge from SNP *rs1372519* located in the *SNCA* locus towards BMI and from *rs2230288*, located in the *GBA* locus, and *rs28399664*, located in the *BCAM* locus, towards exposure to air pollution.

### 4.5 Mendelian Randomization Identifies Causal Impact of Hypertension on PDD

After calibrating the effects of the SNPs from the summary statistics and applying our instrument selection thresholds, only daytime sleep, diabetes, obesity, smoking, hearing loss, and hypertension had SNPs that met the criteria: 37 for daytime sleep, 293 for diabetes, 3 for obesity, 22 for smoking status, 8 for hearing loss, and 463 for hypertension, which makes them suitable for MR analysis. IVW analysis indicated that hypertension increased the risk of developing Parkinson’s disease (PD) dementia, with a causal estimate of 0.223 [se = 0.06, p = 0.0005], and diabetes also presented a risk with a causal estimate of 0.138 [se = 0.03, p = 0.0004]. Daytime sleep did not show a significant risk of dementia due to PD [p = 0.3], nor did obesity [p = 0.87], hearing loss [p = 0.35] or smoking status [p = 0.27]. The detailed results of our MR analysis are found in Supplementary Tables 4 and 5.

MR-Egger sensitivity analysis confirmed a significant causal effect of hypertension on PDD with an estimate of 0.3 [se = 0.19, p = 0.04] but did not reach significance for diabetes [p = 0.22], daytime sleep [p = 0.27], obesity [p = 0.64] or smoking status [p = 0.78]. This may be due to MR-Egger’s lower power compared to IVW analysis due to the inclusion of the intercept term. No comorbidities showed signs of directional pleiotropy, as indicated by insignificant p-values for the MR-Egger intercept. MR-PRESSO analysis indicated horizontal pleiotropy for diabetes [rss = 338.7, p = 0.03], but causal effects remained significant after correction for outliers, with an estimate of 0.18 [se = 0.03, p = 0.000002] and no outlier distortion [p = 0.3]. The global test did not show signs of horizontal pleiotropy for hypertension or daytime sleep. Together, our MR analysis confirmed a potential causal link between hypertension and the risk of dementia from PD. Furthermore, our MR analysis indicates a likely causal impact of type 2 diabetes on the risk of developing PDD.

## 5 Discussion

To our knowledge, our study is the first to explore the predictability of PDD in PwPD and the interplay of various genetic and non-genetic factors in UKB. We developed a machine learning model that predicts PDD in people with Parkinson’s disease (PwPD), achieving an average performance of 0.62 AUC. Although this performance may not be suitable for clinical use, it could be valuable for patient stratification to reduce sample sizes in clinical trials [48]. Additionally, it allowed us to evaluate the contributions of different data modalities, demonstrating that genetic factors had the most significant cumulative impact on the risk of PDD, followed by demographics and comorbidities.

The primary predictor of the model was the polygenic risk score PGS4281, calculated for all-cause dementia using 110 risk variants [32]. In our study, this score alone allowed us to predict PDD with an AUC of 57.5%.

The second most significant predictor was variant *rs769449*, located in the Apolipoprotein E *APOE* locus. Although the APOE *ϵ*4 allele is well recognized as a risk factor for sporadic Alzheimer’s disease (AD) [49], recent research shows that it also impacts *α*-synuclein pathology and its associated toxicity, which are critical aspects of Parkinson’s disease (PD) pathology [50]. As expected, age was among the main predictors in this model, as dementia is generally more prevalent in older people, and PwPD can develop dementia years after motor symptoms appear. However, age alone allowed one only to predict PDD slightly above the change level (AUC 56%), highlighting the need to consider different risk factors in combination.

The variant *rs6859* which is located in the *NECTIN2* locus was among the top predictors and has previously been linked to deterioration of cognitive abilities in adults [51]. We observed a strong influence of sex as a predictor, which is consistent with reported statistics indicating that men are twice as likely as women to develop PDD [52].

Another relevant predictor was body mass index, which had an inverse relation to the probability for PDD. A high body mass index has previously been associated with a protective effect against fast cognitive decline and dementia development in PwPD [53][54][55].

We identified various comorbidities as significant predictors, including depression, anxiety, hypertension, hypercholesterolemia, and excessive daytime sleepiness. Depression and anxiety are recognized non-motor symptoms of Parkinson’s disease (PD) that can precede motor symptoms [56]. Excess daytime sleepiness is more prevalent in patients with PD with dementia compared to those with normal cognitive performance [57]. In addition, both hypertension and hypercholesterolemia are associated with an increased risk of dementia [58][59].

Looking at important genetic factors, we observed variants located in the *APOE* locus, including *rs449647* and *rs7412*. Furthermore, the variant *rs157580*, located in the *TOMM40* locus, and *rs60049679*, located in the *APOC1* locus, which make up the well-known Alzheimer’s disease risk gene cluster, *APOE-TOMM40-APOC1* [60]

A notable predictor was tea intake. A previous MR analysis indicated that green tea consumption may slow PD progression and protect against dementia [61]. In contrast, another MR analysis found that the consumption of more than 13 cups of tea per day was associated with an increased risk of AD dementia [62], possibly due to pesticide residues [63].

We examined interactions between phenotype-related variables and genotype-related features using BN structure learning. The expected interactions included the effect of age on hypertension, daily physical activity, and daytime sleepiness. Unexpected connections were also found, such as between maternal smoking and childhood obesity, possibly reflecting social deprivation. Furthermore, we observed links between sex, smoking status, and anxiety, as the prevalence of smoking and anxiety disorders varies between men and women [64][65]. Some interactions involved variables from different data modalities. For example, the variant *rs1372519* in the *SNCA* locus was associated with body mass index. Although individuals with Parkinson’s disease (PwPD) generally have a lower BMI [66], a direct connection between the *SNCA* mutation and the differences in BMI remains undetermined. Moreover, two edges from *rs2230288* in the *GBA* locus and *rs28399664* in the *BCAM* locus indicated exposure to air pollution. The susceptibility of PwPD to certain gene mutations in regard to air pollution is not well documented, although similar gene-environment interactions have been identified in other diseases, such as chronic obstructive pulmonary disease (COPD) [67].

Lastly, we used MR analysis to examine the causal effects of type 2 diabetes, hypertension, hypercholesterolemia, obesity, and daytime sleepiness on the risk of developing PDD. MR analysis for other comorbidities was not possible, as none of the SNPs obtained from their GWAS summary statistics passed our instrument selection criteria. Our analysis revealed a putative causal effect of hypertension and type 2 diabetes. Studies have shown that elevated blood pressure in middle-aged populations (40-65 years) may be associated with a higher risk of cognitive impairment, while lower blood pressure in an elderly population (≥65) has been associated with a higher prevalence of dementia [68]. Another study has shown that hypertension is related to severe basal ganglia dilated perivascular space (BGdPVS), which in turn is associated with white matter hyperintensities (WMH) and cognitive decline in Parkinson’s disease (PD) [69]. Similarly, previous studies show that type 2 diabetes is associated with faster motor and cognitive impairment in PD [70], and that long-term variability in blood glucose increases the risk of dementia in PD patients [71].

Our study has several limitations, including the reliance on ICD coded reports of PD and PDD in UKB data, which may introduce unknown errors, particularly in distinguishing PDD from other types of dementia, such as Lewy Body Disorder. Furthermore, the generalizability of our findings is affected by the fact that most of the participants were white British, older on average, and predominantly male. Furthermore, the sample size for PD and PDD in UKB is still relatively small compared to the general disease population. We were able to partially address this aspect by investigating the replicability of our machine learning model and the SHAP analysis using data from the PPMI study. However, the number of PDD patients with comorbidities investigated in the UKB was inadequate to replicate our MR analysis.

## 6 Conclusion

In conclusion, to our knowledge, our study is the first to explore the predictability of PDD in PwPD based on data from the UKB population study. We demonstrated that prediction performances on PPMI with newly diagnosed PwPD were comparable, and the impact of individual features on model predictions was similar in both PPMI and UKB. Although prediction performance may overall be insufficient for clinical application, our innovative approach combining machine learning, BN structure learning, and MR analysis has shed light on the complex interactions among genetic predisposition, comorbidities, environment, and lifestyle factors contributing to PDD. In particular, we identified potential causal links between hypertension, diabetes, and PDD, suggesting that the management of blood pressure and glucose levels in the blood in individuals with Parkinson’s might be suitable targets for preventive strategies.

## 7 Acknowledgment

This research has been conducted using the UK Biobank resource under application 67829. Data used in the preparation of this article was obtained from the Parkinson’s Progression Markers Initiative (PPMI) database www.ppmi-info.org/access-dataspecimens/download-data, RRID:SCR 006431. For up-to-date information on the study, visit www.ppmi-info.org. PPMI – a public-private partnership – is funded by the Michael J. Fox Foundation for Parkinson’s Research and funding partners. A list of names of all the PPMI funding partners can be found at www.ppmi-info.org/about-ppmi/who-we-are/study-sponsors/.

## 8 Data Availability

This research has been conducted using the UK Biobank resource under application number 67829. The genetic and phenotype datasets are not publicly available but can be accessed via the UK Biobank data access process. More details are available at http://www.ukbiobank.ac.uk/register-apply/. PPMI data are publicly available from the Parkinson’s Progression Markers Initiative (PPMI) database www.ppmi-info.org/access-data-specimens/download-data, RRID:SCR 006431. For up-to-date information on the study, visit www.ppmi-info.org.

## 9 Code Availability

The underlying code used for model training, mendelian randomization and learning the bayesian networks in this study will be published on GitHub upon acceptance of the paper under the CC BY-NC-ND 4.0 license and can be accessed via https://github.com/mosala777/Predicting-Dementia-in-PwPD

