## Supplementary Figures for "Predicting Dementia in People with Parkinson’s Disease"

**Supplementary Materials**

Mohamed Aborageh^1,∗^,Tom Hähnel^1,2^, Patricia Martinsconde^3^, Jochen Klucken^4^, Holger

Fröhlich^1,4,∗^

^1^Department of Bioinformatics, Fraunhofer Institute for Algorithms and Scientific Computing

(SCAI), 53757 Sankt Augustin, Germany

^2^Department of Neurology, University Hospital and Faculty of Medicine Carl Gustav Carus,

TUD Dresden University of Technology, Dresden, Germany

^3^Luxembourg Centre for Systems Biomedicine (LCSB), University of Luxembourg,

Esch-sur-Alzette, Luxembourg

^4^ Centre Hospitalier de Luxembourg (CHL), Luxembourg

^5^Bonn-Aachen International Center for Information Technology (B-IT), Rheinische

Friedrich-Wilhelms-Universität Bonn, 53115 Bonn, Germany

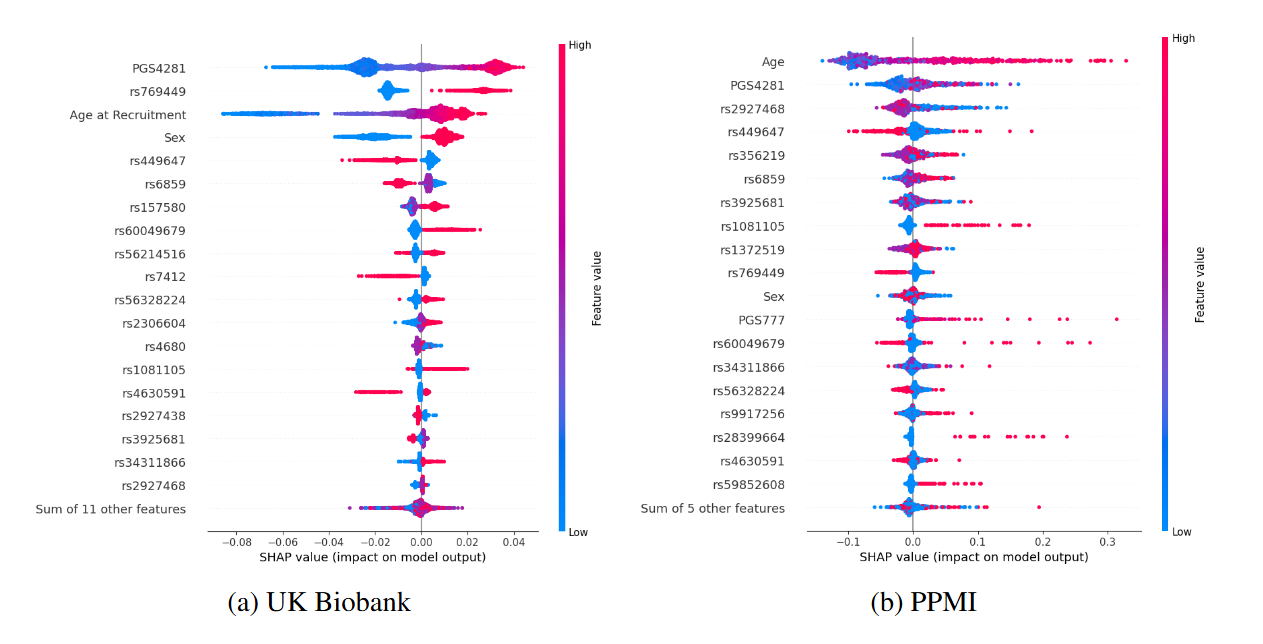

**Supplementary Figure 1**: Comparison of beeswarm plots illustrating the SHAP values for the top predictors identified by the best-performing random forest model on the UK Biobank and PPMI datasets. The x-axis represents the SHAP values, which quantify the contribution of each feature to the model's prediction for a given data point. Positive SHAP values indicate that a feature increases the likelihood of predicting PDD, while negative values suggest a decrease. The y-axis lists the top predictors, ordered by their overall importance, with the most influential features at the top. Each dot represents an individual data point, with its position reflecting the SHAP value and its color indicating the corresponding feature value.

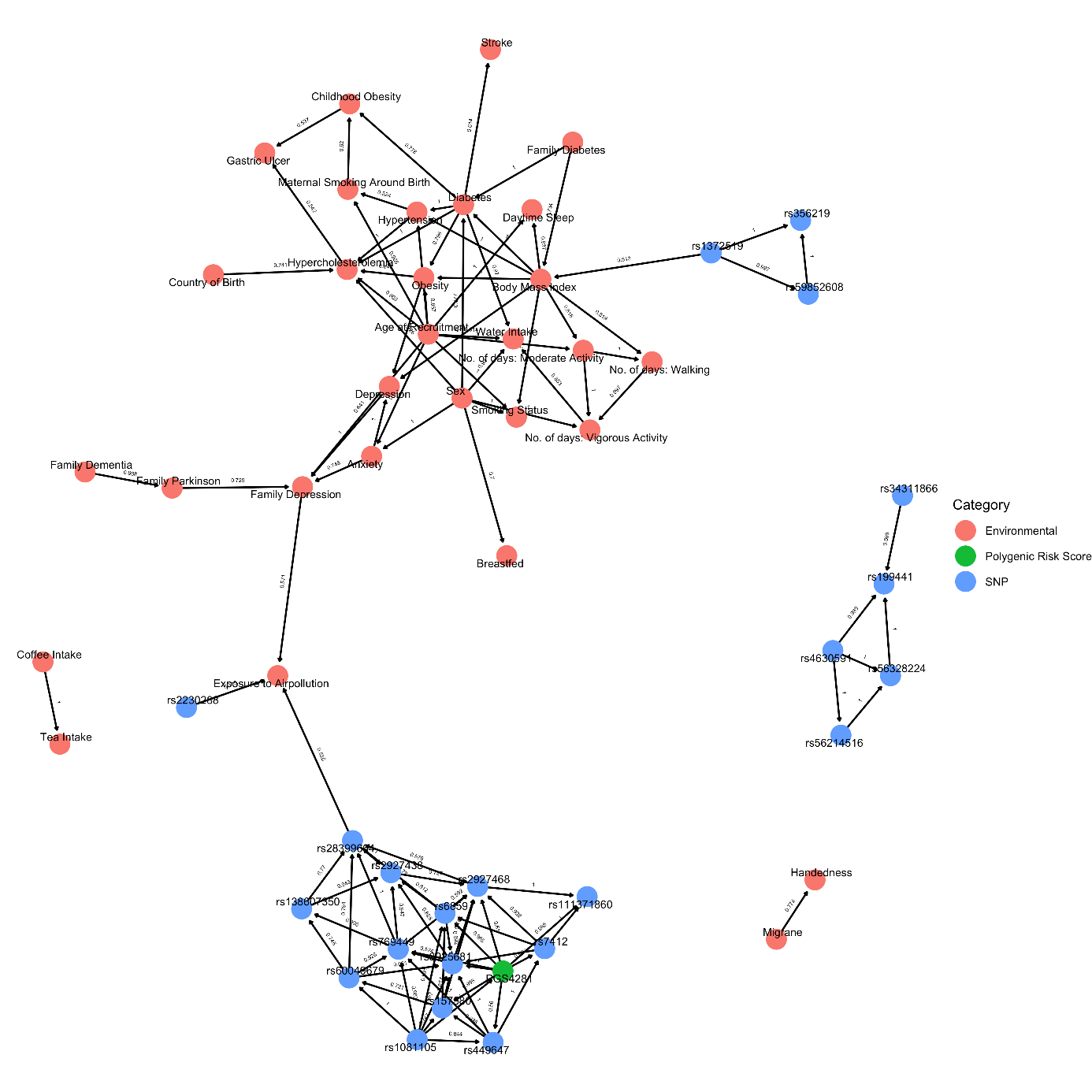

**Supplementary Figure 2**: A graph illustrating the high-confidence edges identified in the Bayesian Network (BN) trained on data from all subjects. The BN was constructed using non-parametric bootstrapping, where random samples were drawn 1,000 times to learn a complete network structure. Edges shown in the graph represent relationships that were consistently identified in at least 50% of all generated networks, indicating high confidence in their presence.

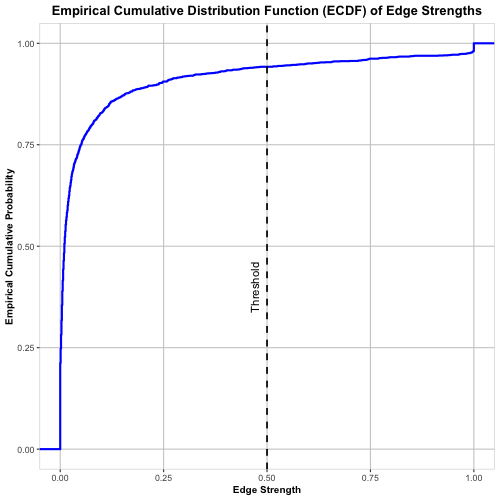

Supplementary figure 3: A figure illustrating the empirical distribution function of the Bayesian network bootstrap frequencies. The x-axis represents edge strength, which quantifies the confidence or weight of edges in the network, ranging from 0 (low strength) to 1 (high strength). The y-axis represents the empirical cumulative probability, showing the proportion of edges with strengths less than or equal to a given value.
